# Resolution of blood RNA signatures fails to discriminate sputum culture status after eight weeks of tuberculosis treatment

**DOI:** 10.1101/2023.11.24.23298983

**Authors:** Claire J Calderwood, Alvaro Sanchez Martinez, James Greenan-Barrett, Blanché Oguti, Jennifer Roe, Rishi Gupta, Adrian R Martineau, Mahdad Noursadeghi

## Abstract

**Background:** There is concerted effort to reduce the burden of 6 months antimicrobial treatment for tuberculosis (TB). Early treatment cessation at 8 weeks is effective for most but incurs increased risk of disease relapse. We tested the hypothesis that blood RNA signatures of TB disease or C-reactive protein (CRP) measurements discriminate microbiological cure after 8 weeks of treatment, as a pre-requisite for a biomarker to stratify risk of relapse.

**Methods:** We identified blood RNA signatures of TB disease or cure by systematic review. We evaluated CRP measurements and blood RNA signatures that could be reproduced in genome-wide transcriptomic data from a previously reported longitudinal dataset in pulmonary TB, spanning samples collected pre-treatment, at 2 and 8 weeks of treatment, and after 2 years of follow up. In our primary analysis, we tested discrimination of sputum culture positivity at 8 weeks by contemporary blood RNA and CRP measurements using area under the receiver operating characteristic curve (AUROC) analysis. In secondary analyses, we tested the relationship between biomarker measurements and time to culture positivity as a surrogate for bacterial load in sputum culture positive cases at 8 weeks, and discrimination of sputum culture status at 8 weeks by biomarker measurements at any other time point.

**Findings:** We evaluated 12 blood RNA signatures. Blood RNA signature scores normalised over time from TB treatment initiation. 11/44 cases with available blood RNA, CRP and sputum culture results, were sputum culture positive at 8 weeks of treatment. None of the 12 blood RNA signature scores tested achieved statistically significant discrimination between sputum culture-positive vs. negative patients at this time point, with AUROC point estimates of 0.48-0.61. CRP achieved the best AUROC of 0.69 (95% confidence interval 0.52-0.87). None of the contemporary biomarker measurements correlated with bacterial load, and no measurements pre-treatment or at 2 weeks discriminated sputum culture status at 8 weeks.

**Interpretation:** The current repertoire of blood RNA signatures of TB and CRP will not provide host response surrogates of microbiological cure to support cessation of TB treatment at 8 weeks. Decoupling of blood transcriptional host-response from the presence of viable bacteria is indicative of subpopulations of *Mycobacterium tuberculosis* able to colonise the respiratory tract without triggering a detectable immune response.

**Research in context:** *Evidence before this study:* We performed a systematic review, using comprehensive terms for “tuberculosis”, “transcriptional” and “biomarker” with no language or date restrictions in Medline on October 4, 2023. Many studies have described normalisation of blood RNA signatures during the course of tuberculosis treatment. Five studies have evaluated blood RNA signatures as a test of microbiological cure after completion of 6 months of treatment. However, there is growing interest in their application as a test of cure to support shortened treatment regimens. The performance of one blood RNA signature has been reported to provide modest discrimination of contemporary sputum culture status at 8 weeks of treatment among HIV co-infected patients with recurrent tuberculosis. We found no reports of whether these findings are generalisable to other blood RNA signatures or to HIV negative patients with their first episode of tuberculosis, who are most likely to be candidates for shortened treatment regimens.

*Added value of this study:* To our knowledge, we provide the first evaluation and comparison of multiple blood RNA signatures of tuberculosis for discrimination of microbiological cure after 8 weeks of tuberculosis therapy among HIV negative patients. 12 previously validated blood RNA signatures of tuberculosis identified by systematic review underwent head-to-head evaluation, alongside blood C-reactive protein measurement as an alternative biomarker of disease, to determine whether they discriminated contemporary sputum culture status after 8 weeks of treatment among 44 HIV negative patients with smear-positive drug-sensitive tuberculosis enrolled to a previously reported randomised controlled trial of adjunctive vitamin D therapy. None of the blood RNA signatures showed statistically significant discrimination of contemporary sputum culture status after 8 weeks tuberculosis treatment, or quantitative relationships with sputum bacterial load among sputum culture positive cases. Importantly, most sputum culture positive cases at this time point, showed contemporary blood RNA signature scores within the normal range.

*Implications of all the available evidence:* Assuming that microbiological cure is a pre-requisite for early cessation of antimicrobial treatment for tuberculosis, the current repertoire of blood RNA signatures of tuberculosis does not provide host response surrogates of microbiological cure to support introduction of 8-week treatment regimens. Therefore, there is a need for further discovery and validation of new biomarkers to support risk stratification for truncated therapy both for research and clinical practice applications. The lack of association between blood transcriptomic signatures and sputum culture status after 8 weeks of treatment suggests the existence of microbial sub-populations that do not trigger a host response. Whether this reflects a state of latency or bacterial persistence in immune privileged compartments requires further investigation.

## Introduction

Standard treatment for drug-sensitive pulmonary tuberculosis (TB) involves at least six months of combination antimicrobial treatment, which exceed 95% cure rates in optimal settings^1^. Clinical trials of antimicrobial treatment regimens have reported over 85% people achieving cure with 3-4 months of treatment^2–6^. These underpinned a novel approach to reduce the treatment burden by using boosted 8-week antimicrobial regimens, followed by extended treatment for persistent disease, monitoring after treatment and retreatment for relapse. This approach achieved equivalent outcomes to standard care at 96 weeks with a 12% non-inferiority margin^7^. However, approximately 20% of people experienced relapse during the follow-up period. Effective approaches to stratify individual-level risk of relapse at the end of shortened TB treatment regimens are required to enable implementation of these approaches in clinical practice.

There has been sustained interest in blood RNA biomarkers of TB as triage tests for symptomatic TB^8–10^, and for pre-symptomatic or incipient TB^11^. Taken together with the observation that their levels can change from the first week of therapy and normalise with treatment^12,13^, this piqued interest in their application to predict clinical outcomes or as a test of cure, with potential to provide greater sensitivity than resolution of symptoms, and overcome the difficulties in obtaining sputum for microbiological testing. A number of studies have reported that these biomarkers discriminate contemporary microbiological treatment failure at the end of standard 6-month treatment regimens^12,14–16^. Only one study has evaluated the performance of an 11-gene (Darboe11) signature to discriminate contemporary microbiological cure before completion of 6 months treatment, as a pre-requisite for their potential utility to stratify risk of relapse following truncated treatment regimens. This showed statistically significant, but modest discrimination of early and late sputum culture conversion after 8 weeks treatment with point estimate for area under the receiver operating characteristic curve (AUROC) of 0.73, but was limited to HIV co-infected individuals with recurrent TB^17^. It is not known whether this finding is generalisable to other gene signatures or to HIV negative patients with their first episode of TB that may be candidates for shortened treatment regimens. We addressed this knowledge gap by testing the hypothesis that blood RNA biomarkers of TB discriminate early microbiological treatment response (8-week sputum culture conversion) among people with pulmonary TB by undertaking a comparative analysis of candidate signatures, identified by systematic review, in a longitudinal clinical trial dataset^18^. We and others have shown that the long-established blood biomarker of inflammation, C-reactive protein (CRP) has comparable performance to blood RNA biomarkers of TB as triage test for disease^19,20^. Therefore, we also evaluated CRP as a test of microbiological cure/response to treatment at 8-weeks alongside our analysis of blood RNA biomarkers.

## Methods

### Study approvals

Blood sampling for transcriptomic analyses performed in the present study was approved by UK National Research Ethics services (reference number: 06/Q0605/83). All subjects provided written informed consent.

### Selection of biomarker signatures

We updated a previous systematic search (on 14/02/2023) to identify candidate blood RNA biomarkers, using the same databases and search strategy as described previously^8^ (Supplementary Methods, Supplementary File 1). All records identified by the original and updated searches were sifted in duplicate (ASM, BO, CC) to reflect updated inclusion criteria. In the current review, we included articles that described discovery of at least one concise whole blood mRNA signature for the purpose of assessing the response to TB treatment, as well as those for diagnosis of active or incipient TB. Only signatures which had been validated in at least one separate dataset and were fully reproducible through a published calculation or regression equation were included. Signatures comprised of multiple genes are referred to as the surname of the first author of the corresponding publication, suffixed by the number of constituent genes.

### Study cohort and biomarker measurements

Genome-wide blood transcriptomic data were obtained using cDNA microarrays, as previously described^21^, in a subset of 46 people (Supplementary Table 1) with smear and culture-positive drug-sensitive pulmonary TB enrolled to a randomised controlled trial of adjunctive vitamin D therapy (AdjuVIT)^18^. We included a random sample from participants in whom sputum culture results were available at eight weeks post-treatment, and longitudinal blood RNA samples were available pre-treatment and any of two weeks, eight weeks or two years post-treatment (Supplementary Figure 1). Sputum cultures and CRP measurements at eight weeks post-treatment were performed as part of the study protocol in the original trial. CRP levels after eight weeks treatment were previously reported, stratified by treatment arm and participant vitamin D receptor genotype^22^, but not previously related to 8-week sputum culture status.

### Data Analysis

Analyses were performed using R (version 3.6.1). Blood RNA signature scores were calculated using the authors’ described methods. Blood RNA signatures with more than 20% of constituent genes missing, because they were not represented among protein-coding genes within the microarray dataset, were excluded (Supplementary Table 1). Signature scores were scaled by Z-scores standardised to the mean and standard deviation of measurements from a subset of participants with sustained cure two years after treatment completion (N=31). Receiver operating characteristic curve (ROC) analysis using the pRoc package evaluated two-class discrimination, with pairwise comparisons of ROC curves performed using the DeLong method^23^. Sensitivity and specificity were calculated at the maximum Youden index of the ROC curve, and pre-specified thresholds of a Z-score of 2 (Z2) for blood RNA signatures and 5 mg/L for CRP. The primary analysis focussed on biomarker discrimination area under the ROC curve (AUROC) of sputum culture status at 8 weeks of treatment. In addition, we undertook secondary analyses to test the hypothesis that biomarker levels at 8 weeks correlated with quantitative bacterial load in sputum culture positive individuals at 8 weeks of treatment, and to evaluate discrimination of 8 week sputum culture status by biomarker measurements in the preceding time points (pre-treatment, 2 weeks and 4 weeks).

The inclusion criteria for selection of gene signatures in the present systematic review excluded the Darboe11 signature because this required re-derivation by a support vector machine learning model. However, we recently showed that the average expression of these genes performs as well as original SVM derived signature^20^. Since this signature had been previously reported to discriminate 8-week sputum culture conversion, we included the average signature score as a supplementary analysis in the present study.

### Data availability

The microarray data presented in this study will be made available in an open-access public repository at the time of peer-reviewed publication.

### Statistical power

The sample size was determined by the number of cases from the AdjuVIT trial for which blood RNA, CRP and sputum culture results were available. We evaluated the power to identify statistically significant (p<0.05) discrimination of sputum culture positive and culture negative cases from total sample size N=44 stratified by ratio of positive:negative control cases and AUROC (PASS2022 software) (Supplementary Figure 2).

## Results

### Resolution of candidate blood RNA biomarkers of TB with curative treatment

1133 publications identified by systematic review yielded 38 studies and 49 blood RNA signatures of TB that met the criteria for inclusion. 37 of the 49 blood RNA signatures were excluded from further analysis either as they required model recreation (n=16), because there were signatures of lesser accuracy within the same study (n=12), due to the signature assessing an incorrect indication (n=3), or due to missing genes (n=6). 12 blood RNA signatures were included in downstream analyses. (Supplementary Figure 3, Supplementary Table 2, Supplementary File 1).

Blood RNA signature scores normalised over time from TB treatment initiation (Figure 1). Consistent with their identification as biomarkers of active TB disease, most of these showed high classification accuracy for discriminating between pre-treatment measurements and 2-year post-treatment measurements (AUROC range 0.67–0.99, Supplementary Figure 4, Supplementary Table 3). At the prespecified Z2 threshold which approximates specificity to 98% by virtue of standardising biomarker scores using the distribution of 2-year post-treatment scores, the blood RNA signatures achieved sensitivity point estimates of 20-98% for identification of TB disease represented by the pre-treatment samples. A statistically significant reduction in the distribution of blood RNA signature scores compared to pre-treatment baseline measurements, was evident at 2 weeks for one single gene signature (ADM), and for all other signatures by 8 weeks treatment.

**Figure 1.**
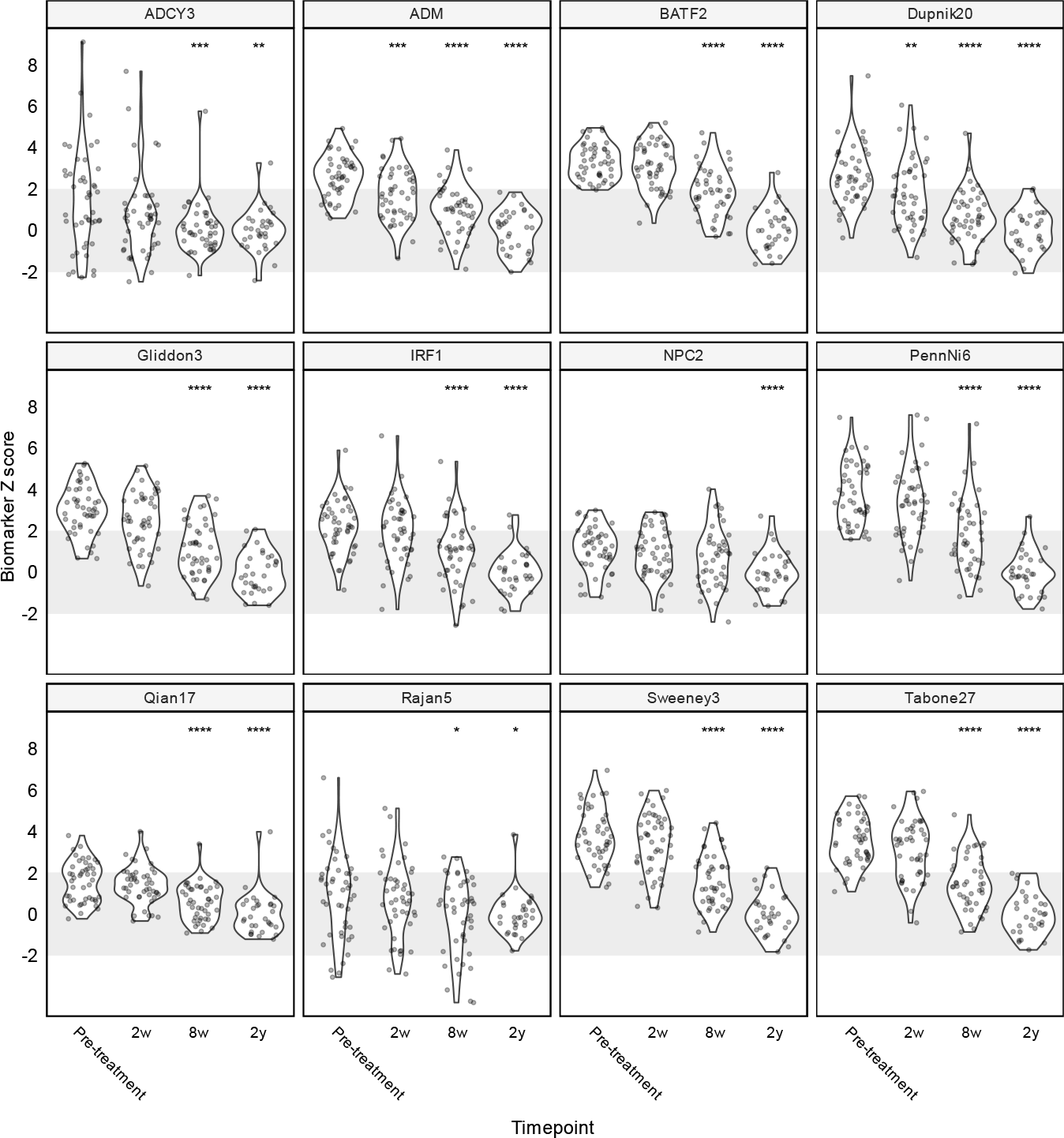
Longitudinal blood RNA signature scores by time from start of TB treatment. Blood RNA signature scores standardised to the distribution of 2 year post-treatment samples are shown pre-treatment, and 2 weeks, 8 weeks and 2 years (2y) from initiation of treatment. 2 week, 8 week and 2 year time points were compared to pre-treatment by Wilcoxon Rank Sum test. *p values for change from baseline to timepoint indicated with * p <0.05, ** p <0.01 *** p <0.001 and **** p <0.0001. Grey shaded area indicates Z-score +/-2 (normalised to 2-year values).

### Biomarker discrimination of sputum culture status at 8 weeks post-treatment

11 of 44 patients with blood RNA data available at 8 weeks of treatment had contemporary Mtb culture positive sputum (Supplementary Figure 1). In the primary analysis, this case mix provided 80% power to discriminate culture positive and negative cases with AUROC >0.75 (Supplementary Figure 2). None of the 12 blood RNA signature scores tested achieved statistically significant discrimination between patients by sputum culture status at this time point, with AUROC point estimates of 0.48-0.61. CRP achieved the best AUROC of 0.69 (95% CI 0.52-0.87) (Figure 2, Supplementary Figure 5, Supplementary Table 4). Importantly, using the Z2 threshold as the upper limit of normal range derived from patients with sustained cure 2 years post-treatment, we found most sputum culture-positive individuals at 8 weeks with blood RNA and CRP measurements in the normal range, indicative of the presence of viable bacteria without a discernible host response.

**Figure 2.**
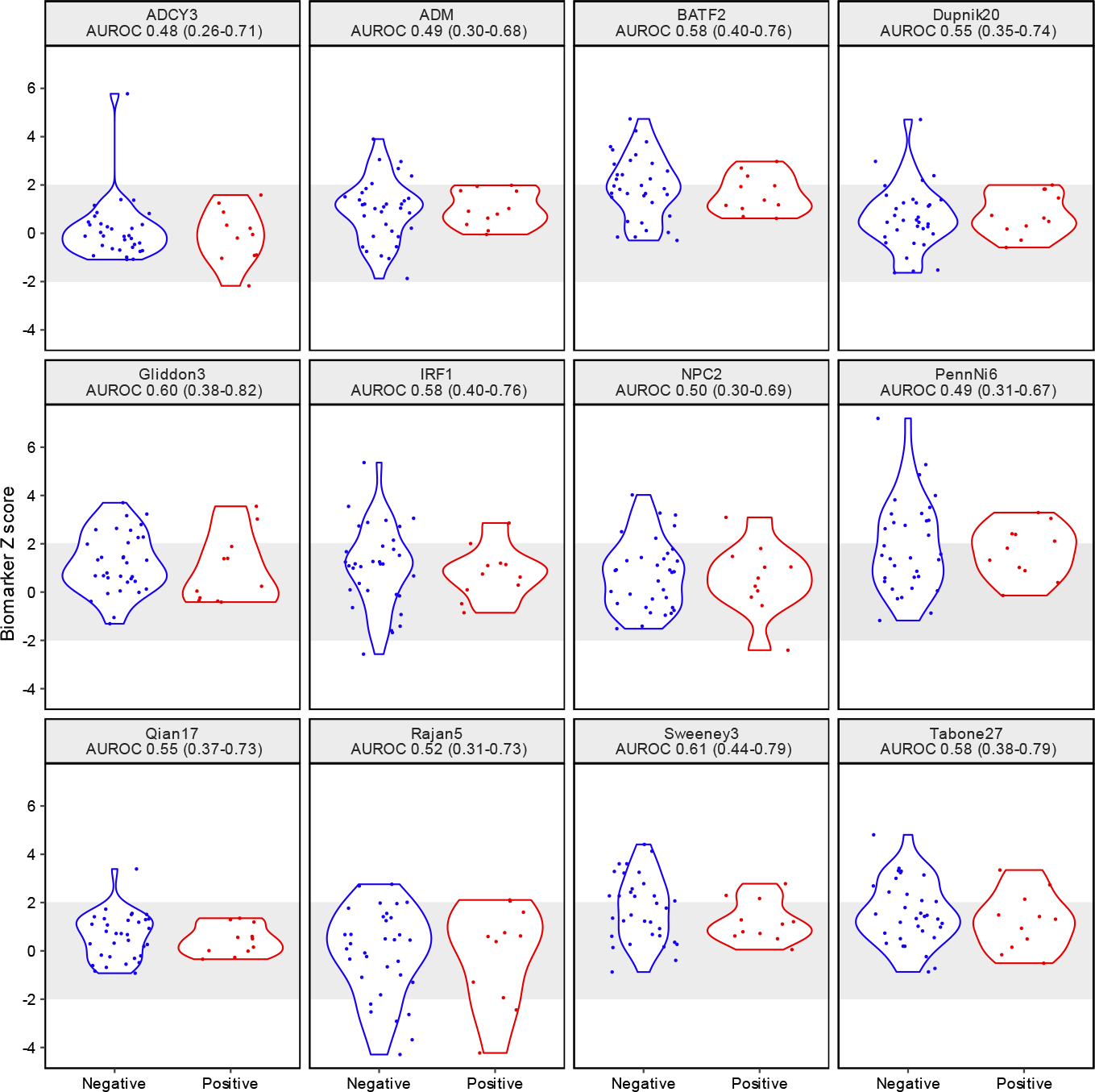
Blood RNA signature scores and sputum culture status at 8 weeks. Distributions of blood RNA signature scores at 8 weeks of treatment for tuberculosis, stratified by contemporary sputum culture status, showing AUROC (±95% confidence intervals) for each signature.

We performed three exploratory secondary analyses. First, we reasoned that the magnitude of host-response as reflected by biomarker levels may be quantitatively related to bacterial load. However, among patients who were sputum culture positive at 8 weeks, none of the contemporary biomarker measurements showed a statistically significant correlation with time to culture positivity as a surrogate for bacterial load (Figure 3). Second, we tested the hypotheses that the host response pre-treatment, early resolution of host responses at 2 weeks post-treatment initiation or the rate of resolution of host responses on treatment over the first 8 weeks may discriminate sputum culture status at 8 weeks. None of these were evident in our analysis (Supplementary Figure 6-7). Third, we tested the performance of the Darboe11 signature that had previously been reported to provide some discrimination between sputum culture positive and negative cases after 8 weeks treatment among HIV co-infected people with pulmonary tuberculosis^17^. Herein, we used the average expression of the genes in this signature which we found to be equivalent to the original support vector machine model as a biomarker of active TB^20^. The performance of the modified Darboe11 signature was comparable to all other signatures in our analysis. This signature normalised with time on treatment, but did not discriminate sputum culture status or relate to bacterial load at 8 weeks of treatment (Supplementary Figure 8).

**Figure 3.**
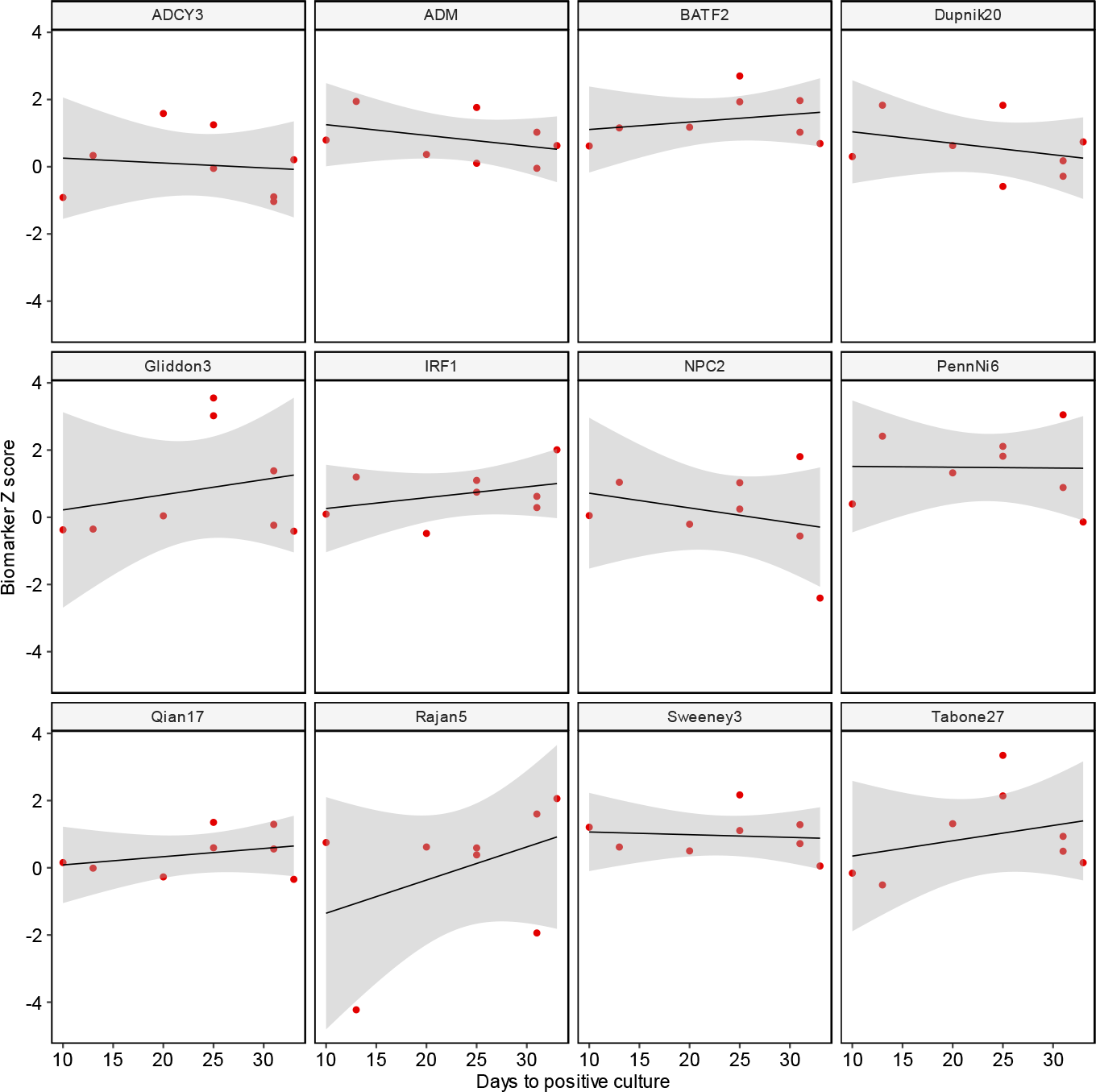
Relationship between blood RNA signature and bacterial load at 8weeks. Scatter plot of blood RNA signature scores with sputum culture time (days) to positivity as a surrogate of bacterial load at 8 weeks of TB treatment showing individual data points and linear model (with 95% confidence limits) for sputum culture positive cases.

## Discussion

To the best of our knowledge, we report the first evaluation of the resolution of blood RNA biomarkers of TB and CRP as a test of sputum culture conversion to reflect microbiological cure following 8 weeks of antimicrobial treatment for TB. Even though elevated pre-treatment biomarker levels reduced to levels within the pre-specified normal range by 8 weeks of treatment, none of the blood RNA biomarkers showed statistically significant discrimination of contemporary sputum culture status at this time point. CRP achieved marginal albeit statistically significant discrimination. In addition, we found no relationship between contemporary bacterial load and biomarker levels as continuous variables, and no relationship between biomarker levels at earlier time points or their response to treatment with 8-week sputum culture conversion. These results suggest that the current repertoire of blood RNA signatures of TB and CRP will not provide host response surrogates of microbiological cure.

Our findings also provide novel biological insights suggesting that the host response may be ‘decoupled’ from the clearance of *Mtb* from the lung. Importantly, given blood RNA biomarkers can detect pre-symptomatic incipient TB^8^, our data provide the most direct evidence to date for Mtb colonisation of the respiratory tract without a discernible host response. This interpretation is further supported by our recent report of detection of Mtb DNA in circulating haematopoietic stem cells without perturbation of the blood transcriptome providing bona fide evidence of latent infection^24^. A possible explanation is that these blood RNA signatures reflect a host response to some Mtb subpopulations, rather than all Mtb, and that these populations may have differential rates of clearance on starting treatment. Since sputum culture conversion only measures the bacilli that can be expectorated, these two tests may not be measuring the same Mtb populations. Testing blood biomarkers against culture conversion, as a proxy for disease activity, may therefore never achieve a test of microbiological cure. Importantly, sputum culture conversion at 8 weeks has not faithfully predicted long term treatment outcomes in clinical trials^25^. Mycobacterial genetic material has been detected in pulmonary samples long after TB cure, suggesting that residual bacillary material is not exclusively associated with disease^26^. Therefore, it may be that blood RNA biomarkers at the time of treatment cessation using truncated regimens can predict long term disease relapse even if they cannot predict contemporary microbial sterilisation.

Strengths of this study include the use of a comprehensive systematic review to select transcriptional signatures for head-to-head analysis alongside that of CRP. Quantitative bacterial load data from sputum cultures also enabled us to test for continuous relationships between host response and bacterial load. The small sample size of study population is a limitation but does not detract from the observation that normalisation of blood RNA signatures at 8 weeks did not provide an adequate test of Mtb clearance from sputum at this timepoint. Some of the reconstructed signatures were incomplete because they contained genes, miRNAs or pseudogenes that were missing in the AdjuVIT dataset. Nonetheless, they achieved comparable accuracies to the complete signatures. Some blood RNA signatures were excluded completely because we were not able to reconstruct them adequately in our data set. However, previous analysis that shows co-correlation of these signatures^8^, suggests that our findings are likely to be generalisable to other blood RNA signature of active TB.

In view of the limitations of sputum culture conversion as an end point, future evaluation of these signatures should be undertaken in treatment shortening trials to assess their ability to discriminate patients with and without relapse after the end of treatment. Such studies should continue to incorporate genome-wide transcriptional profiling to support discovery of novel signatures for this application and head-to-head analysis of different signatures as they emerge.

## Author contributions

MN and ARM conceived the study. CC, ASM, BO performed the systematic review. JR performed the blood transcriptional profiling. CC, ASM, RKG performed the data analysis. CC and MN wrote the paper with input from all the authors.

