## Supplementary methods and data for "Resolution of blood RNA signatures fails to discriminate sputum culture status after eight weeks of tuberculosis treatment"

..

### Supplementary methods

#### *Search strategy for blood RNA signatures*

We searched Medline and Embase on 14/02/2023, restricting to articles published after the date of our previous search (15/04/2019), using the same search terms as previously. The search included comprehensive terms for 'biomarkers' (any of Biomarkers/Diagnostic Tests, Routine/"Predictive Value of Tests"/diagnostic test*.mp./biomarker*.mp./ppv.mp./npv.mp./sensitivit*.mp./specificit*.mp./signature*.mp.) AND 'tuberculosis (any of exp TUBERCULOSIS/tuberculosis.mp./exp MYCOBACTERIUM TUBERCULOSIS/tb.mp.) AND 'transcriptome' (any of RNA/Transcriptome//rna.mp./transcript*.mp./gene expression.mp./Gene Expression Profiling/RNA, Messenger/Transcription, Genetic/Gene Expression/) AND 'blood' (any of blood/lood.mp.), limited to human data. We consolidated the search by hand-searching reference lists of relevant review articles. New search results were merged with the results of the previous search and screened by two independent reviewers.

#### *Eligibility criteria for candidate signatures*

We included concise whole blood mRNA signatures discovered with a primary objective of either diagnosis of active or incipient TB, compared to controls who were either deemed healthy, or had latent TB infection, or, assessing the response to TB treatment, either as longitudinal change over time or with reference to another marker of treatment response (e.g. culture conversion, treatment failure or relapse). We defined 'concise' as signatures that used a defined approach to feature selection to reduce multidimensionality and the number of constituent genes, thus leading to biomarkers that may be more amenable to clinical translation, as previously. Other inclusion criteria were that the publications included validation of performance in cases/controls not included in discovery of the signature, availability of the gene names that comprise the signature, availability of the equation or modelling approach. We excluded gene signature models that required de novo derivation of the model in the original data set. Where genes required for a signature were missing from the analysis (AdjuVIT) dataset, we created a 'restricted' signature in the original dataset and compared the results to those obtained using the 'full' signature. If we were unable to reproduce statistically comparable results to those reported, the signature was excluded. Where multiple signatures were discovered for the same intended purpose and from the same training dataset within individual publications, we included the signature with greatest accuracy (as defined by the area under the receiver operating characteristic curve (AUROC) in the validation data), or if accuracy was equivalent, we included the most parsimonious signature.

#### *Data extraction*

Data were extracted using a pre-defined template by one reviewer and independently checked by a second.

#### *Missing gene symbols*

We reviewed gene symbols in g:PROFILER (https://biit.cs.ut.ee/gprofiler/convert) to identify all gene symbol and map to AGILENT microarray probe ID available in the analysis dataset. Signatures with greater than 20% genes missing were excluded from the downstream analysis.

### Supplementary Table 1

***Summary characteristics of cases.***

| | **Included in analysis** | **Excluded from analysis** | | | | |
|---|---|---|---|---|---|---|
| **Characteristic** | **Sputum culture converted at 8 weeks** | **Sputum culture converted at 8 weeks** | | | | |
| | | **No**, N = 11[1] | **Yes**, N = 35[1] | **No**, N = 21[1] | **Yes**, N = 53[1] | **Not known**, N = 38[1] |
| Age | | 36 (31, 42) | 29 (24, 36) | 34 (29, 40) | 30 (25, 41) | 37 (27, 53) |
| Gender | Female | 1 (9.1%) | 7 (20%) | 4 (19%) | 14 (26%) | 14 (37%) |
| | Male | 10 (91%) | 28 (80%) | 17 (81%) | 39 (74%) | 24 (63%) |
| BMI | | 20.21 (19.77, 21.12) | 20.15 (18.48, 21.48) | 18.93 (17.63, 20.42) | 19.92 (17.96, 22.26) | 19.09 (17.31, 21.88) |
| Days to positive culture[2] | | 11.0 (8.0, 13.5) | 13.0 (9.0, 21.0) | 9.0 (5.5, 11.0) | 10.0 (8.0, 15.0) | 11.0 (8.0, 13.4) |
| Ethnicity[3] | BLAF | 2 (18%) | 11 (31%) | 5 (24%) | 24 (45%) | 9 (24%) |
| | EASIA | 1 (9.1%) | 3 (8.6%) | 0 (0%) | 3 (5.7%) | 2 (5.3% |
| | EURAM | 4 (36%) | 3 (8.6%) | 11 (52%) | 7 (13%) | 14 (37%) |
| | SASIA | 4 (36%) | 18 (51%) | 5 (24%) | 16 (30%) | 13 (34%) |
| Educated >18 years | | 2 (18%) | 20 (57%) | 8 (38%) | 26 (49%) | 14 (37%) |
| CXR cavitation | Absent | 5 (45%) | 13 (37%) | 6 (29%) | 26 (49%) | 23 (61%) |
| | Present | 6 (55%) | 22 (63%) | 15 (71%) | 27 (51%) | 15 (39%) |
| Sputum smear grade[4] | ≤1+ | 5 (45%) | 23 (66%) | 3 (14%) | 25 (47%) | 17 (52%) |
| | >2+ | 6 (55%) | 12 (34%) | 18 (86%) | 28 (53%) | 16 (48%) |
| CRP / mg/L[5] | | 42 (26, 82) | 50 (28, 66) | 73 (50, 100) | 58 (38, 86) | 56 (29, 76) |

*[1]Median (IQR) or Frequency (%). [2]Days to positive culture available on 43 people included in present analysis and 84 people not included. [3]In addition, two people of MIDEAST and one person of mixed ethnicity were in the cohort not included in the current analysis. [4] 5 people (all not included in the analysis) were missing baseline smear status. [5] 26 people (1 included in analysis and 25 not included) had missing CRP. BLAF= Black African, EASIA=East Asian, EURAM= European/American, SASIA=South Asian.*

**Supplementary Figure 1**

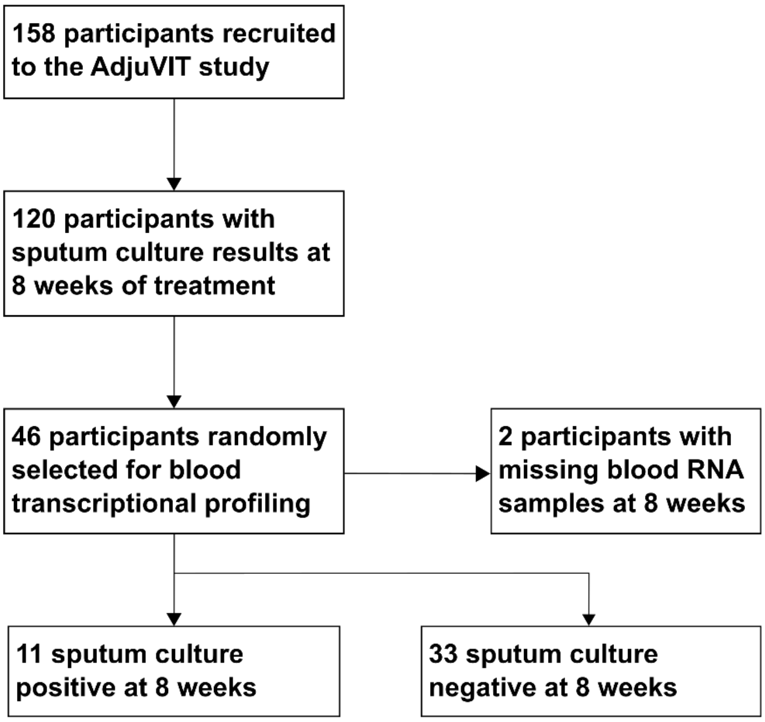

***Consort diagram for samples included in the present analysis.***

**Supplementary Figure 2**

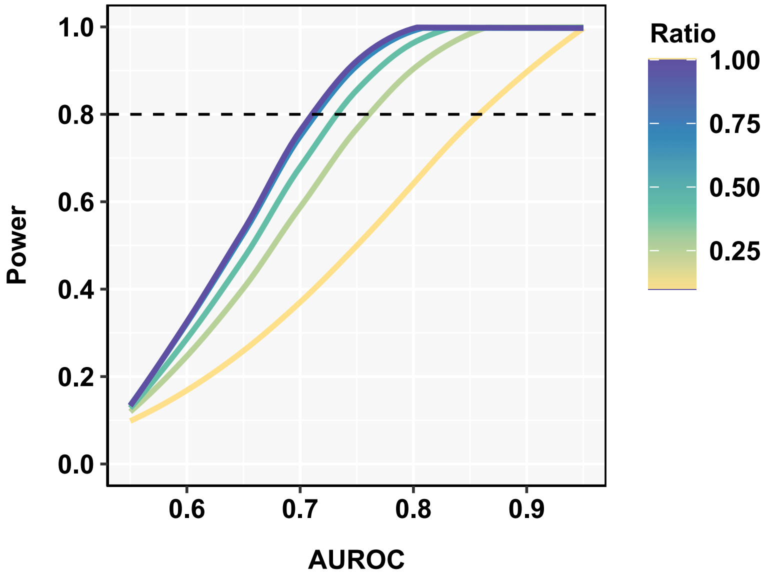

***Statistical power of sample size***

Power to identify statistically significant ($p<0.05$) discrimination of sputum culture positive and culture negative cases from total sample size N=44 stratified by ratio positive:negative cases and AUROC for which biomarker data were available at 8 weeks (PASS2022 software).

### Supplementary Figure 3

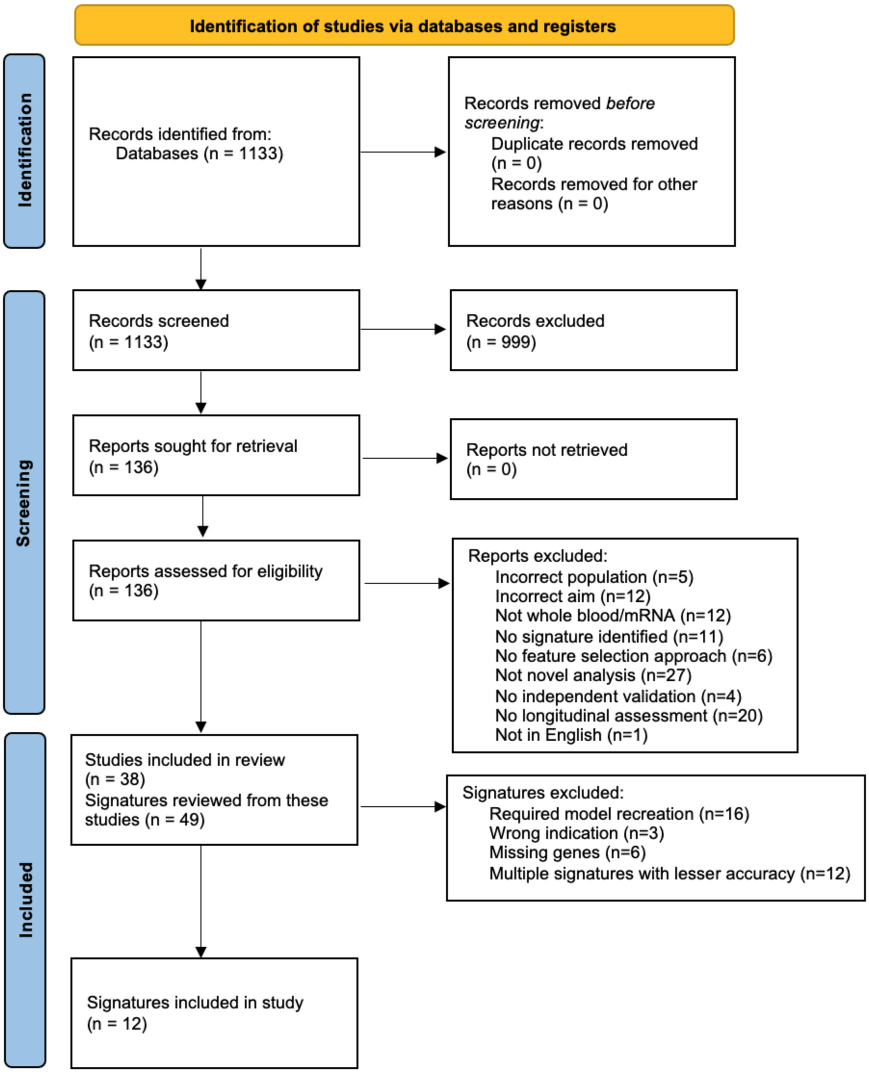

***PRISMA diagram for selection of blood RNA signatures of TB by systematic review.***

**Supplementary Table 2**

***Composition of blood RNA signatures included***

| **Blood RNA signature** | **Total genes** | **N missing** | **% missing** | **Score calculation** |
|---|---|---|---|---|
| ADCY3 | 1 | 0 | 0 | Single gene score |
| ADM | 1 | 0 | 0 | Single gene score |
| BATF2 | 1 | 0 | 0 | Single gene score |
| Dupnik20 | 20 | 2 | 10 | Sum(NAIP,ANXA3,SIPA1L2,CREB5,CCM2,SRPK1,SOCS3,PGLYRP1,LIMK2,CR1,CACNA1E,GYG1,DYSF,RAB20,PFKFB3,MAPK14,ENTPD1,PHTF1,WDFY3,GK) |
| Gliddon3 | 3 | 0 | 0 | Sum(FCGR1B,C1QB,ZNF296) |
| IRF1 | 1 | 0 | 0 | Single gene score |
| NPC2 | 1 | 0 | 0 | Single gene score |
| PennNi6 | 6 | 0 | 0 | mean(GBP2,FCGR1B,SERPING1)-mean(TUBGCP6,TRMT2A,SDR39U1) |
| Qian17 | 17 | 3 | 18 | Sum(MT2A,HERC5,PSMB9,MOV10,PLAAT4,ISG20,WDFY1,GBP1,OAS1,PARP9,IFITM3,GCH1,SAMD9,STAT1,SCARB2,HLADMA,HLADMB) |
| Rajan5 | 5 | 0 | 0 | Sum(GBP6,ACTA2,MTRF1L,GYG1,RABL2A) |
| Sweeney3 | 3 | 0 | 0 | (GBP5+DUSP3)/2)-KLF2 |
| Tabone27 | 27 | 2 | 7 | Mean(CDCP1,C1QC,GBP6,C1QB,PRTN3,BAK1,ICAM1,SCARF1,IGF2BP3,SEPTIN4,SOCS3,LIMK1,GRAMD1C,GRIN3A,IL22,NRN1,CASP5,FCGR1BP,FCGR1CP,FCGR1A,GBP5,SDC3,PDCD1LG2,TNFSF11,P2RY14) |
| Darboe | 11 | 1 | 9 | Mean(BATF2,ETV7,FCGR1A, GBP1,GBP2,GBP5,SCARF1,SERPING1,STAT1,TAP1) |

**Supplementary Figure 4**

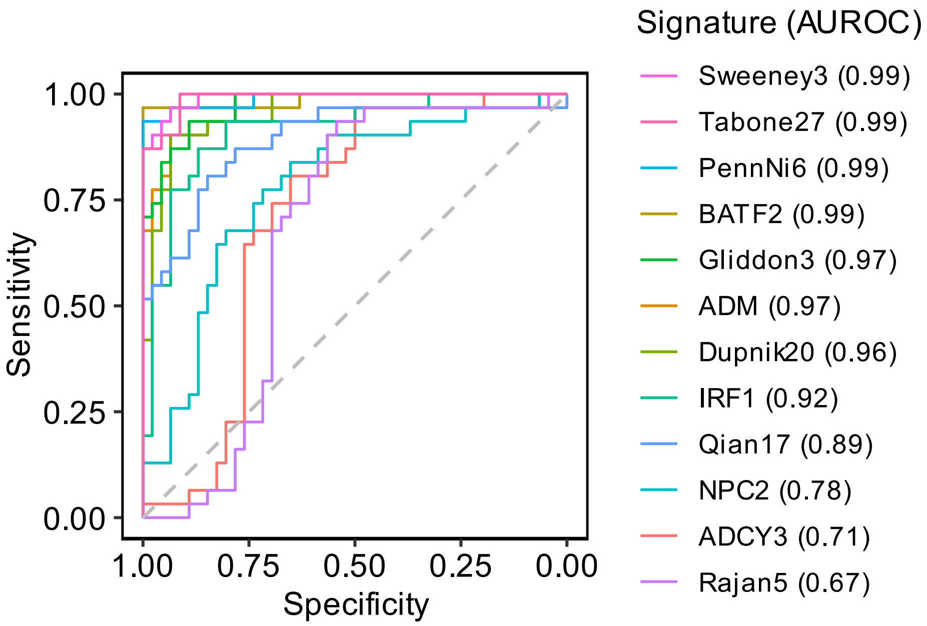

***Blood RNA signature discrimination of pre-treatment and 2-year post treatment samples from AdjuVIT study.***

Receiver operating characteristic curves for discrimination of pre-treatment samples and 2-year post-treatment samples from the AdjuVITs tidy by blood RNA signatures shown (with AUROC point estimates).

### Supplementary Table 3

| Signature | Reported AUROC in original publication | Observed AUROC in present data | Youden index | | Z > 2 | |
|---|---|---|---|---|---|---|
| | | | **Sensitivity** | **Specificity** | **Sensitivity** | **Specificity** |
| Sweeney3 | 0.90 (0.85-0.95) | 0.99 (0.98-1.00) | 0.97 (0.84-1.00) | 0.93 (0.82-0.98) | 0.91 (0.80-0.97) | 0.97 (0.84-1.00) |
| Tabone27 | 0.96 (0.92-1.00) | 0.99 (0.98-1.00) | 1.00 (0.89-1.00) | 0.91 (0.80-0.97) | 0.91 (0.80-0.97) | 1.00 (0.89-1.00) |
| PennNi6 | 0.94 (0.86-1.00) | 0.99 (0.97-1.00) | 0.94 (0.79-0.98) | 1.00 (0.92-1.00) | 0.85 (0.72-0.92) | 0.97 (0.84-1.00) |
| BATF2* | 0.99 (0.96-1.00) | 0.99 (0.96-1.00) | 0.97 (0.84-1.00) | 1.00 (0.92-1.00) | 0.98 (0.89-1.00) | 0.97 (0.84-1.00) |
| Gliddon3 | 0.97 (0.93-1.00) | 0.97 (0.94-1.00) | 0.94 (0.79-0.98) | 0.89 (0.77-0.95) | 0.78 (0.64-0.88) | 0.94 (0.79-0.98) |
| ADM | 0.90 | 0.97 (0.94-1.00) | 0.90 (0.75-0.97) | 0.93 (0.82-0.98) | 0.72 (0.57-0.83) | 1.00 (0.89-1.00) |
| Dupnik20 | NA | 0.96 (0.92-1.00) | 0.90 (0.75-0.97) | 0.93 (0.82-0.98) | 0.70 (0.55-0.81) | 0.97 (0.84-1.00) |
| IRF1 | 0.80 | 0.92 (0.85-0.98) | 0.87 (0.71-0.95) | 0.87 (0.74-0.94) | 0.61 (0.46-0.74) | 0.94 (0.79-0.98) |
| Qian17 | 0.99 (0.98-1.00) | 0.89 (0.81-0.97) | 0.81 (0.64-0.91) | 0.85 (0.72-0.92) | 0.35 (0.23-0.49) | 0.97 (0.84-1.00) |
| NPC2 | 0.94 (0.81-1.00) | 0.78 (0.67-0.89) | 0.77 (0.60-0.89) | 0.72 (0.57-0.83) | 0.20 (0.11-0.33) | 0.97 (0.84-1.00) |
| ADCY3 | 0.88 (0.83-0.93) | 0.71 (0.59-0.83) | 0.97 (0.84-1.00) | 0.50 (0.36-0.64) | 0.41 (0.28-0.56) | 0.97 (0.84-1.00) |
| Rajan5 | 0.87 (0.72-0.98) | 0.67 (0.54-0.80) | 0.94 (0.79-0.98) | 0.54 (0.40-0.68) | 0.24 (0.14-0.38) | 0.97 (0.84-1.00) |

***Performance metrics of blood RNA signatures for discrimination of pre-treatment and 2-year post treatment samples from AdjuVIT study.***

Reported AUROC to discriminate active TB from healthy controls for individual blood RNA signatures are compared to observed AUROC for blood RNA signatures discrimination of pre-treatment and 2-year post treatment samples from AdjuVIT study. Sensitivity and specificity metrics are shown for the Youden Index of the AUROC curve, and using blood RNA signature Z score threshold of >2 (standardised against the distribution of blood RNA signature scores from the 2-year post treatment samples). Point estimates and 95% confidence intervals are shown for each metric.

….

### Supplementary Figure 5

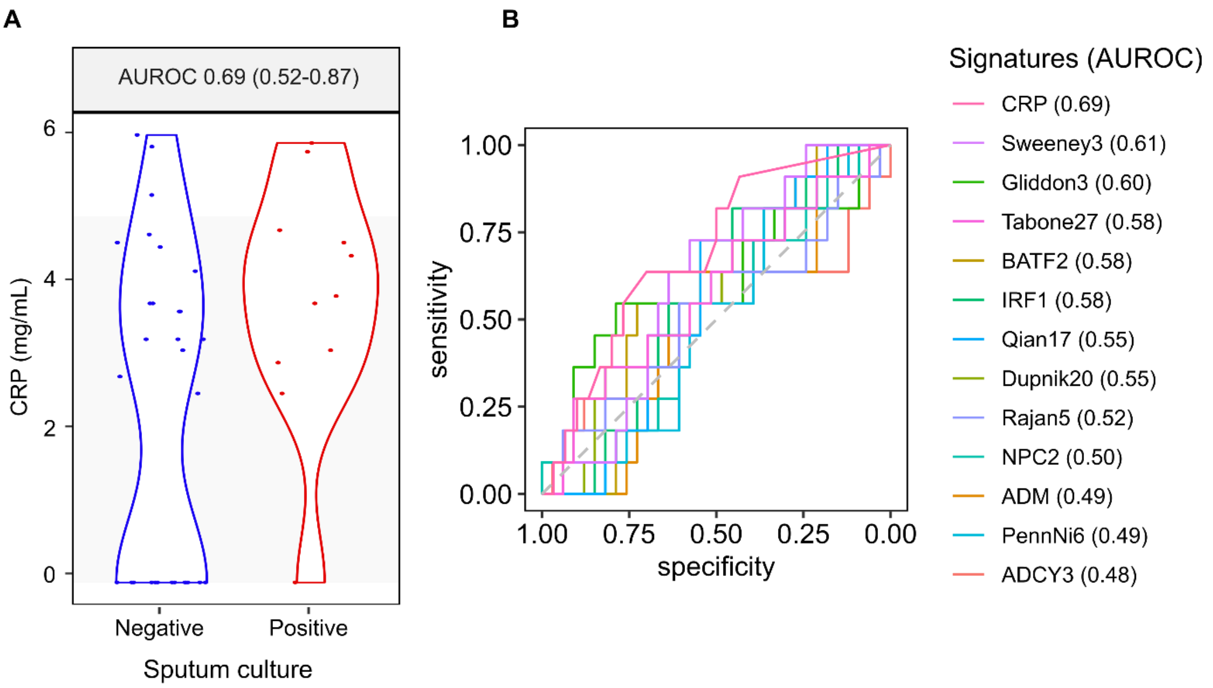

***Biomarker discrimination of contemporary sputum culture result at 8 weeks TB treatment.***

**(A)** Blood C-reactive protein (CRP) measurements stratified by contemporary sputum culture result at 8 weeks of treatment (showing AUROC point estimate and 95% confidence interval for between group discrimination). **(B)** Receiver operating characteristic curve analysis for blood RNA signatures and CRP discrimination of contemporary sputum culture positive and negative cases at 8 weeks TB treatment, showing AUROC point estimates for each biomarker.

### Supplementary Table 4

| | | Max Youden Index | | Z > 2 | |
|---|---|---|---|---|---|
| **Signature** | **AUROC** | **Sensitivity** | **Specificity** | **Sensitivity** | **Specificity** |
| Sweeney3 | 0.61 (0.44-0.79) | 0.73 (0.43-0.90) | 0.58 (0.41-0.73) | 0.73 (0.43-0.90) | 0.45 (0.30-0.62) |
| Gliddon3 | 0.60 (0.38-0.82) | 0.55 (0.28-0.79) | 0.79 (0.62-0.89) | 0.82 (0.52-0.95) | 0.30 (0.17-0.47) |
| Tabone27 | 0.58 (0.38-0.79) | 0.36 (0.15-0.65) | 0.82 (0.66-0.91) | 0.73 (0.43-0.90) | 0.36 (0.22-0.53) |
| BATF2 | 0.58 (0.40-0.76) | 0.55 (0.28-0.79) | 0.73 (0.56-0.85) | 0.73 (0.43-0.90) | 0.42 (0.27-0.59) |
| IRF1 | 0.58 (0.40-0.76) | 0.73 (0.43-0.90) | 0.55 (0.38-0.70) | 0.82 (0.52-0.95) | 0.24 (0.13-0.41) |
| Qian17 | 0.55 (0.37-0.73) | 0.73 (0.43-0.90) | 0.55 (0.38-0.70) | 1.00 (0.74-1.00) | 0.03 (0.00-0.15) |
| Dupnik20 | 0.55 (0.35-0.74) | 0.36 (0.15-0.65) | 0.82 (0.66-0.91) | 0.00 (0.00-0.26) | 0.88 (0.73-0.95) |
| Rajan5 | 0.52 (0.31-0.73) | 0.64 (0.35-0.85) | 0.52 (0.35-0.67) | 0.18 (0.05-0.48) | 0.91 (0.76-0.97) |
| NPC2 | 0.50 (0.30-0.69) | 0.73 (0.43-0.90) | 0.39 (0.25-0.56) | 0.91 (0.62-1.00) | 0.18 (0.09-0.34) |
| ADM | 0.49 (0.30-0.68) | 1.00 (0.74-1.00) | 0.18 (0.09-0.34) | 1.00 (0.74-1.00) | 0.18 (0.09-0.34) |
| PennNi6 | 0.49 (0.31-0.67) | 0.82 (0.52-0.95) | 0.36 (0.22-0.53) | 0.45 (0.21-0.72) | 0.58 (0.41-0.73) |
| ADCY3 | 0.48 (0.26-0.71) | 0.27 (0.10-0.57) | 0.88 (0.73-0.95) | 0.00 (0.00-0.26) | 0.97 (0.85-1.00) |
| CRP | 0.69 (0.52-0.87) | 0.91 (0.62-1.00) | 0.43 (0.27-0.61) | 0.91 (0.62-1.00) | 0.43 (0.27-0.61) |

***Performance metrics of blood RNA signatures for discrimination of sputum culture positive and negative cases at 8 weeks TB treatment.***

Observed AUROC for blood RNA signatures discrimination of sputum culture positive and negative cases at 8 weeks TB treatment from AdjuVIT study. Sensitivity and specificity metrics are shown for the Youden Index of the AUROC curve, and using blood RNA signature Z score threshold of >2 (standardised against the distribution of blood RNA signature scores from the 2-year post treatment samples). Point estimates and 95% confidence intervals are shown for each metric.

### Supplementary Figure 6

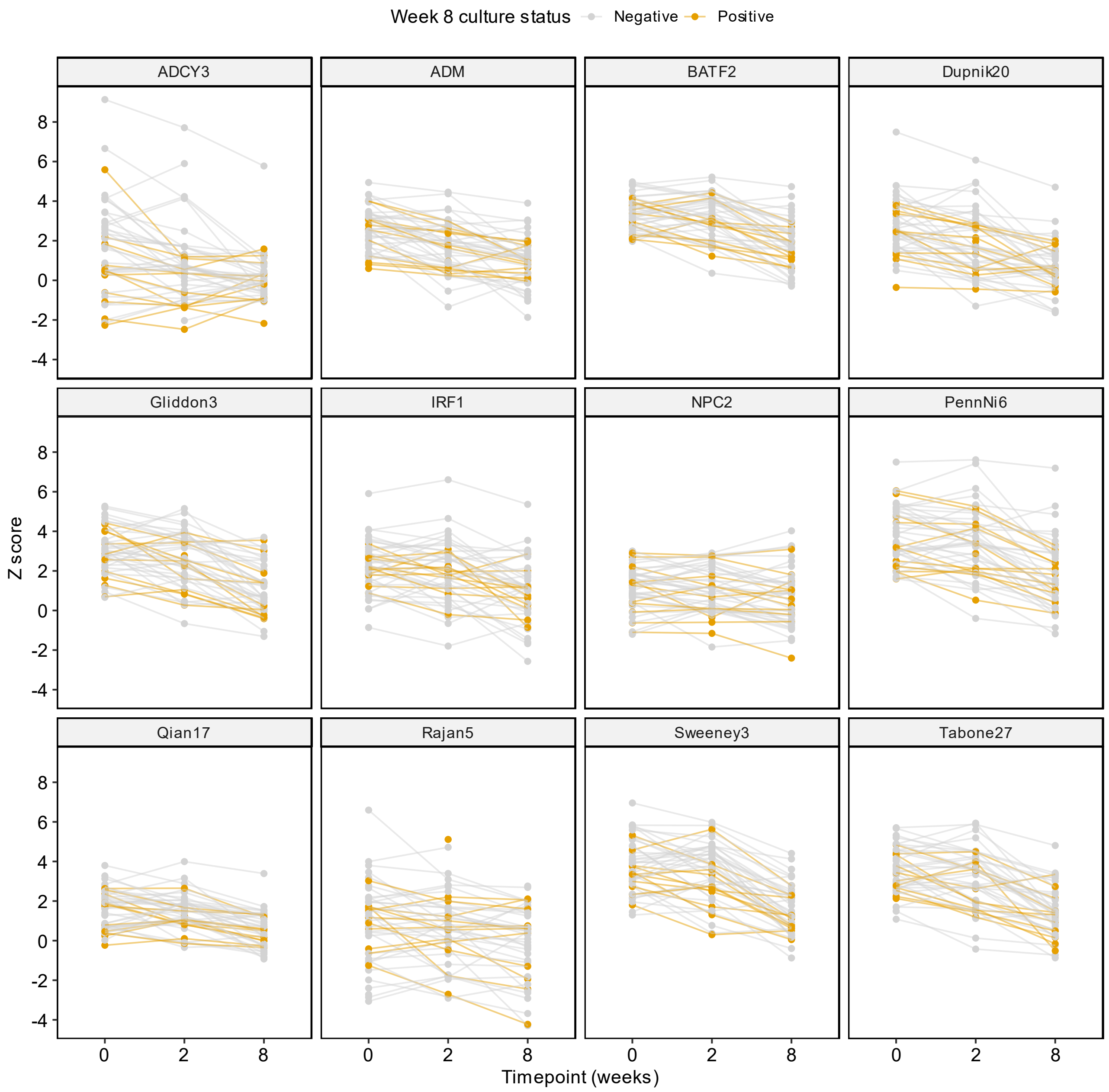

***Longitudinal blood RNA signature scores stratified by sputum culture status at 8 weeks TB treatment.***

Longitudinal blood RNA signature Z scores (standardised against the distribution of blood RNA signature scores from the 2-year post treatment samples) are shown pre-treatment and 2 and 8 weeks TB treatment at for individual participants stratified by sputum culture status 8 weeks of TB treatment.

**Supplementary Figure 7**

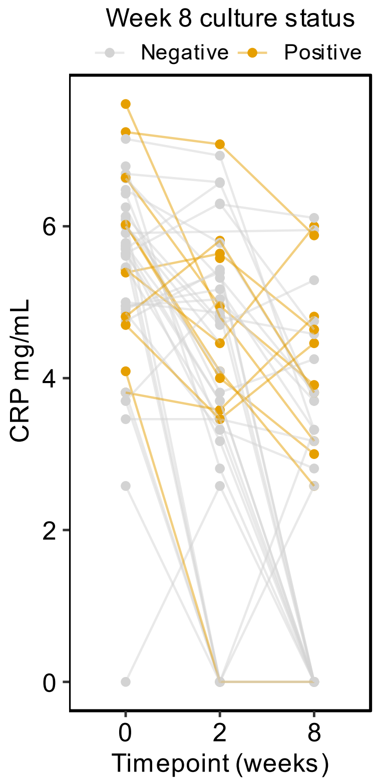

***Longitudinal CRP measurements stratified by sputum culture status at 8 weeks TB treatment.***

Longitudinal C-reactive protein measurements are shown pre-treatment and 2 and 8 weeks TB treatment at for individual participants stratified by sputum culture status 8 weeks of TB treatment.

**Supplementary Figure 8**

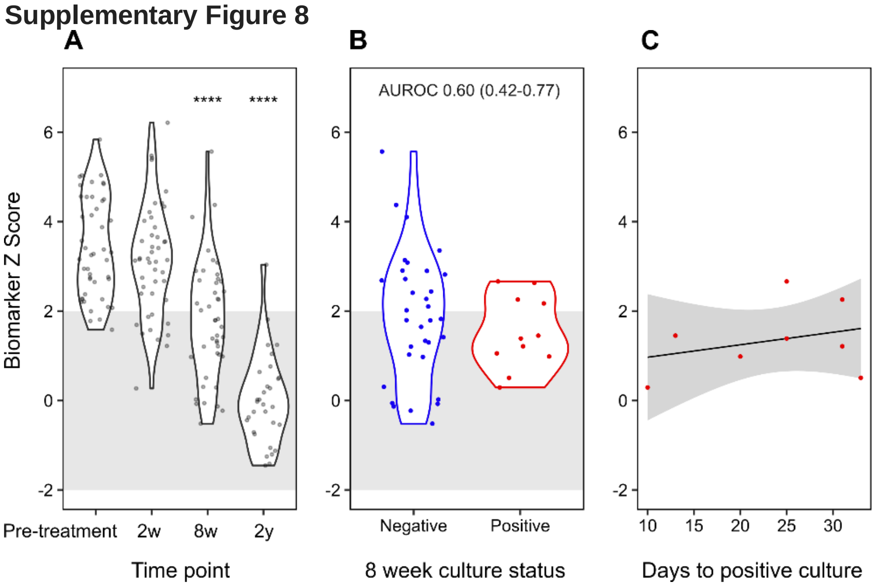

***Performance of modified Darboe11 signature***

**(A)** Signature scores standardised to the distribution of 2 year post-treatment samples are shown pre-treatment, and 2 weeks, 8 weeks and 2 years (2y) from initiation of treatment. 2 week, 8 week and 2 year time points were compared to pre-treatment by Wilcoxon Rank Sum test. *p values for change from baseline to timepoint indicated with * p <0.05, ** p <0.01 *** p <0.001 and **** p <0.0001. Grey shaded area indicates Z-score +/-2 (normalised to 2-year values). **(B)** Distributions of signature scores at 8 weeks of treatment for tuberculosis, stratified by contemporary sputum culture status, showing AUROC (±95% confidence intervals) for each signature. **(C)** Scatter plot of signature scores with sputum culture time (days) to positivity as a surrogate of bacterial load at 8 weeks of TB treatment showing individual data points and linear model (with 95% confidence limits) for sputum culture positive cases.
